# Infectiousness of places: The impact of human settlement and activity space in the transmission of COVID-19

**DOI:** 10.1101/2021.09.02.21263012

**Authors:** Lun Liu, Hui Wang, Zhu Zhang, Weiyi Zhang, Shengsheng Zhuang, Tingmiao Lv, Chi On Chio, Yifan Wang, Daori Na, Chuchang Tang, On Ieng Ao-Ieong

## Abstract

Places are fundamental factors in the spread of epidemics, as they are where people agglomerate and interact. This paper explores how different types of places—activity spaces at micro-level and human settlements at macro-level—impact the transmission of infections using evidences from COVID-19. We examine eleven types of activity spaces and find heterogeneous impacts across countries, yet we also find that non-essential activity spaces tend to have larger impacts than essential ones. Contrary to common beliefs, settlement size and density are not positively associated with reproduction numbers. Further, the impacts of closing activity spaces vary with settlement types and are consistently lower in larger settlements in all sample countries, suggesting more complex pattern of virus transmission in large settlements. This work takes first steps in systematically evaluating the epistemological risks of places at multiple scales, which contributes to knowledge in urban resilience, health and livability.

**Teaser:** Activity spaces and human settlement characteristics impact the spread of epidemics in multiple ways and should be considered in policy making.

## Introduction

Humans continue to migrate to large, dense urban settlements in the past century. The consequent growth of cities brings benefits such as economies of scale and knowledge spillovers, but also increases the vulnerability of human society to risks related to people’s agglomeration and interaction such as infectious disease, pollution and crime (*1, 2*), for which the on-going outbreak of COVID-19 is a prominent example. In tackling these risks, places are important aspects, as the human interactions giving rise to various risks are all associated with certain physical places. To understand the role of different types of places in the spread of risks is important for targeted policy making to contain the risks and enhance the resilience of cities.

In terms of places’ epistemological risks, the COVID-19 pandemic provides worldwide data with natural experiments to investigate the spread of infectious disease at different types of places. Place characteristics at multiple scales could have an impact. At the macro-level, for instance, dense settlements lead to physical proximity among residents which is likely to generate more contacts, and large settlements connect more people—both might increase the dissemination of diseases (*3-5*). At the micro-level, different activity spaces, such as restaurants, museums, sports venues, are likely to be associated with different risks of disease spread, affected by the socioeconomic interactions at these locations.

Despite of a large number of researches on the spread of COVID-19, the potentially varying transmission risks at different places are still not systematically investigated. Macro-scale place characteristics are rarely examined and evidences are mixed (*3, 4, 6*). For micro-scale places, the most relevant body of research is those on the efficacy of non-pharmaceutical interventions on COVID-19 which include the closure of various activity spaces (*7-17*). However, these works mostly use broad categories when estimating the impact of activity spaces, such as “non-essential businesses”, “venues” or “lock down”, which can involve a number of distinct types of spaces (*7, 10, 14, 18, 19*). Since the closure of any activity space could affect the daily life of certain groups, the effect estimates on broad categories are insufficient for governments to make cost-effective intervention policies. As COVID-19 persists and countries have to lock down repeatedly, it is critical that we understand which types of settlements and activity spaces need more rigorous interventions than other, thus refining policies in the on-going COVID-19 and similar crisis in the future.

In this work, we take the case of COVID-19 and investigate how different types of macro-level settlement characteristics and micro-level activity spaces impact the spread of infections, as well as how they interact. We examine two basic characteristics of settlements—population size and density—which have been found to affect many social quantities (*5, 20*), and eleven types of activity spaces commonly included in government interventions, which are schools, childcare centers, offices, non-essential retails, restaurants, bars, entertainment venues, cultural venues, religious venues, indoor sports venues and outdoor sports grounds (detailed descriptions in Table S1). To perform the analysis, we combine data from a variety of sources including COVID-19 infection case data from government data portals and open data repositories, government intervention data from national and state-level government websites, and socioeconomic characteristics of settlements from various official statistics (see Section S1 for details). Four countries from different continents which are strongly hit by the pandemic are chosen as study cases, which are Japan in Asia, the United Kingdom in Europe, the United States in North America and Brazil in South America. We take settlements with population above 100,000 as samples, since many smaller settlements do have enough cases to derive reliable estimates of instantaneous reproduction number, which is the outcome of concern in our analysis. We use data from the first pandemic wave, that is, from March to August 2020, since there could be more uncertainties confounding the analysis in later periods of the pandemic including the so-called “lockdown fatigue”, virus variants, and vaccination (*9*).

Ideally, we would like to take continuous built-up areas as units of analysis, which can be considered as individual settlements. However, continuous built-up areas usually do not overlap with administrative boundaries, based on which infection cases and other statistics are usually published. Given the constraint, we choose the administrative or statistical units which are most similar to continuous built-up areas and where infection case data are available as the spatial units in the analysis, that are, prefectures of Japan, local authority districts of the United Kingdom, metropolitan statistical areas of the United States and municipalities of Brazil (detailed explanations on the choice of spatial units in Section S1.1-1.4). Note that the prefectures of Japan are larger than the other spatial units and usually contain more than one continuous built-up area, however, infection case data can only be consistently acquired at this level in Japan (*21*). Nonetheless, we prove that the results are not likely to be affected by the issue (Section S3.3). The spatial units in each country with population above 100,000 are taken as sample, leading to 45 spatial units in Japan, 234 in the United Kingdom, 308 in the United States and 319 in Brazil after cleaning missing data.

Our methodology is based on an econometric approach called difference-in-differences (DiD), which is widely used in examining causal relationship in social processes (*22*). We first estimate the causal impacts of activity space closures on the course of the epidemic, which can also be interpreted as the risks of virus transmission associated with respective spaces. This is implemented by modelling the relationship between instantaneous reproduction numbers (*R*_*t*_) in the spatial units and the corresponding status of activity spaces, controlling for other government interventions including stay-at-home-orders and gathering bans. The DiD method estimates causal impact by subtracting the course of *R*_*t*_ in spatial units where a certain type of activity space get closed or reopened with the course of *R*_*t*_ in spatial units where the same type of activity space remain unchanged, given that *R*_*t*_ in the two groups should move in parallel trend absence of the change. By subtracting the trends, the method can rule out the impact of common behavioral changes shared by all spatial units such as increased self-protection, which could otherwise be falsely attributed to activity spaces thus inflate the estimates (*10*). We estimate separate models on each country to allow for heterogeneous impacts of activity spaces across countries, which might be influenced by factors including the lifestyle, culture, urban form and building design.

The fact that governments might close or reopen multiple types of activity spaces together poses obstacles for identifying the impact of individual types of activity spaces (*14*). But as time went, there were more timing differences, especially in the reopening stage, to facilitate more nuanced analysis. Correlation analysis shows that the Kendall’s correlation coefficients between the status of activity spaces are mostly smaller than 0.8 in our study period (Fig. 1). We merge activity space closures with correlation coefficients larger than 0.95 as one intervention, after which there are at least 180 unit-day differences between any pair of interventions in Japan (due to only 45 spatial units) and 699 in the other countries. We further verify that the estimated impacts are not sensitive to removing intervention variables, suggesting manageable collinearity (Section S3.2).

**Fig. 1.**
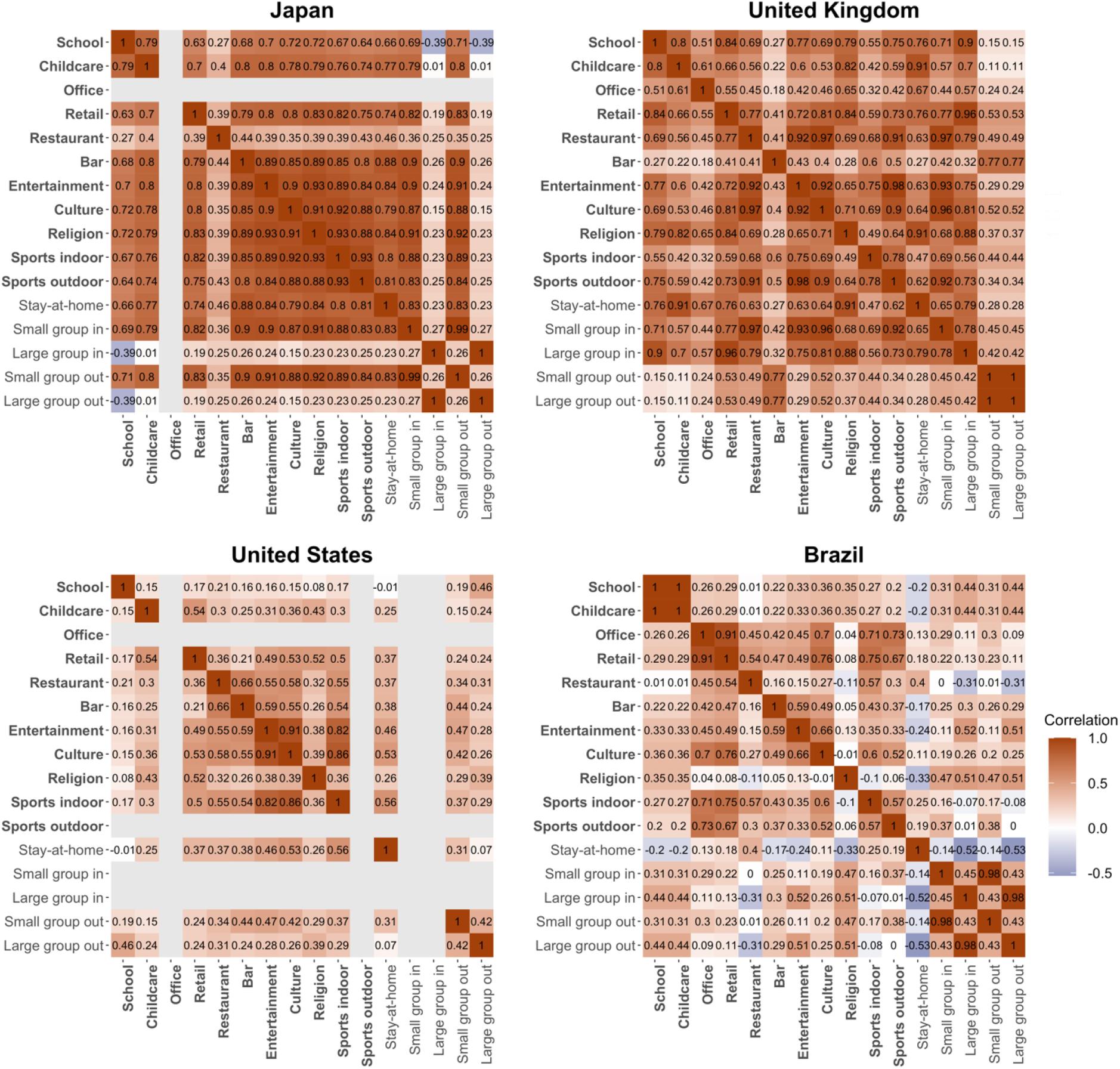
Correlation between the status of activity spaces and other government interventions. The matrices show pairwise Kendall’s correlation coefficients between the status of activity spaces and other government interventions across the spatial units in each country during the study period. Bold texts indicate the activity spaces and regular texts are the other government interventions that we control for. Grey cells indicate missing data. The correlation coefficients are estimated based on samples excluding unit-day observations where the coefficient of variance for *R*_*t*_ estimate is larger than 0.3 (suggesting unreliable estimates).

The DiD estimation is implemented through a two-way fixed effect model with fixed effects of days and spatial units, which is a widely used modelling method to implement DiD analysis (*23*). The fixed effects of spatial units can be interpreted as the intrinsic reproduction number in the spatial units absence of any voluntary or compulsory behavioral changes, based on which we estimate the impacts of settlement size and density while controlling for socioeconomic characteristics of settlements. In addition to measuring the independent impacts of micro-level activity spaces and macro-level settlement characteristics on infections, we also examine how the impacts of activity space closures interact with settlement size and density. This is implemented by repeating the DiD analysis on separate samples of relatively large and small, and high-and low-density spatial units and then comparing the effect sizes of activity space closures. More details on the specification of the models and sensitivity tests can be found in Materials and Methods and Section S3.

## Results

### Impact of micro-level activity spaces

We first estimate the causal impact of closing eleven types of activity spaces on *R*_*t*_ by fitting a two-way fixed effect model on each country’s data, controlling for other government interventions including gathering bans and stay-at-home orders (Fig. 2, full model results in Table S2). The estimated impacts vary across countries, which could stem from varying profiles of visitors to each type of activity space in different countries, varying behaviors in the activity spaces, as well as varying physical conditions of relevant spaces. The activity space closures that show a statistically significant impact on reducing *R*_*t*_ in each country are: entertainment venues (53%, 4 ∼ 77%) in Japan; restaurants and cultural venues (combined with indoor gathering bans whose effect is inseparable, 25%, 5 ∼ 41%) and indoor sports venues (43%, 13 ∼ 63%) in the United Kingdom; entertainment venues (17%, 1 ∼ 31%) in the United States; and non-essential retails (20%, 9 ∼ 31%) and indoor sports venues (36%, 27 ∼ 43%) in Brazil. The percentage reductions in *R*_*t*_ are transformed from direct model outputs (as shown in Fig. 2) as 1-*e*^*x*^, where *x* denotes the direct outputs. These estimates can also be interpreted as the proportions of total infections related to the respective activity spaces, which may either happen inside these places or on the way travelling to these places.

**Fig. 2.**
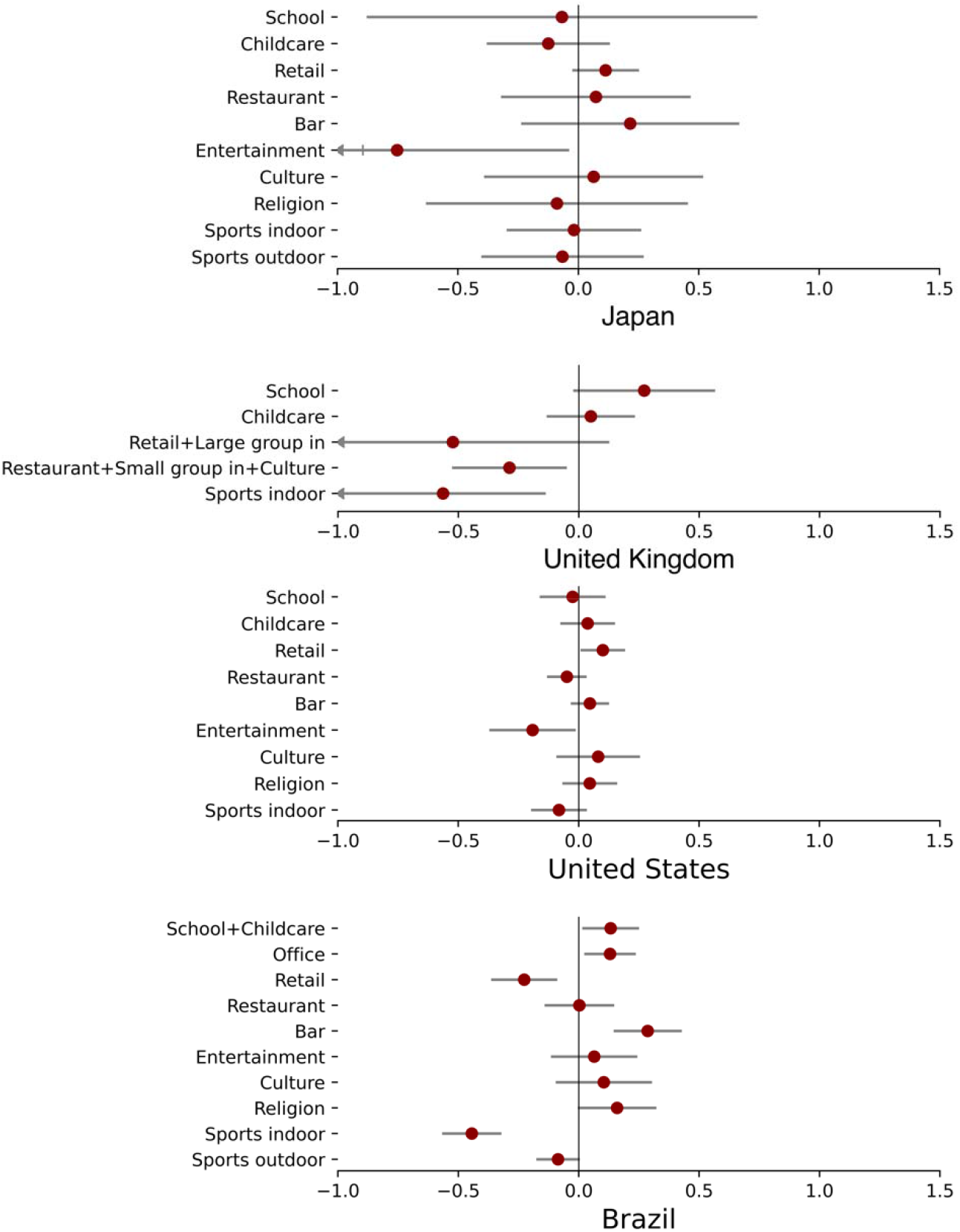
Estimated impacts of closing individual types of activity spaces. The numbers are direct model outputs on the relationship between activity space status (0, 0.5 or 1) and *Δlog(R*_*t*_). Full results are presented in Table S2.

Most of the activity space closures satisfy the parallel trend assumption, meaning that the estimates are not biased by potentially different pre-trends of *R*_*t*_ in areas that close or reopen an activity space and those that do not (detailed methodology and results of the parallel trend test in Section S2.2 and Table S3). The estimates are also generally robust to a number of alternative settings in the analysis, including withholding spatial units from the sample and increasing or decreasing confounding variables in the model, suggesting that they are not likely to be affected by individual influential spatial units and the correlation among variables (detailed methodology and results in Section S3.1 and S3.2).

Considering that governments often need to devise strategies on closing a set of activity spaces in an epidemic, we further estimate the combined effects of multiple activity spaces. We compute the effects and uncertainties of closing all possible combinations of activity spaces in each country based on the modelling results in the last step. The full results are provided on this project’s Github repository https://github.com/lunliu454/infect_place for readers to explore. Here we present the maximum reduction in *R*_*t*_ that can be achieved by closing a given number of types of activity spaces (Fig. 3). Our analysis suggests that the largest reductions in *R*_*t*_ are achieved by closing two to six types of activity spaces, while more closures do not further bring reproduction numbers down. Governments could resort to this kind of analysis when making cost-effective intervention strategies.

**Fig. 3.**
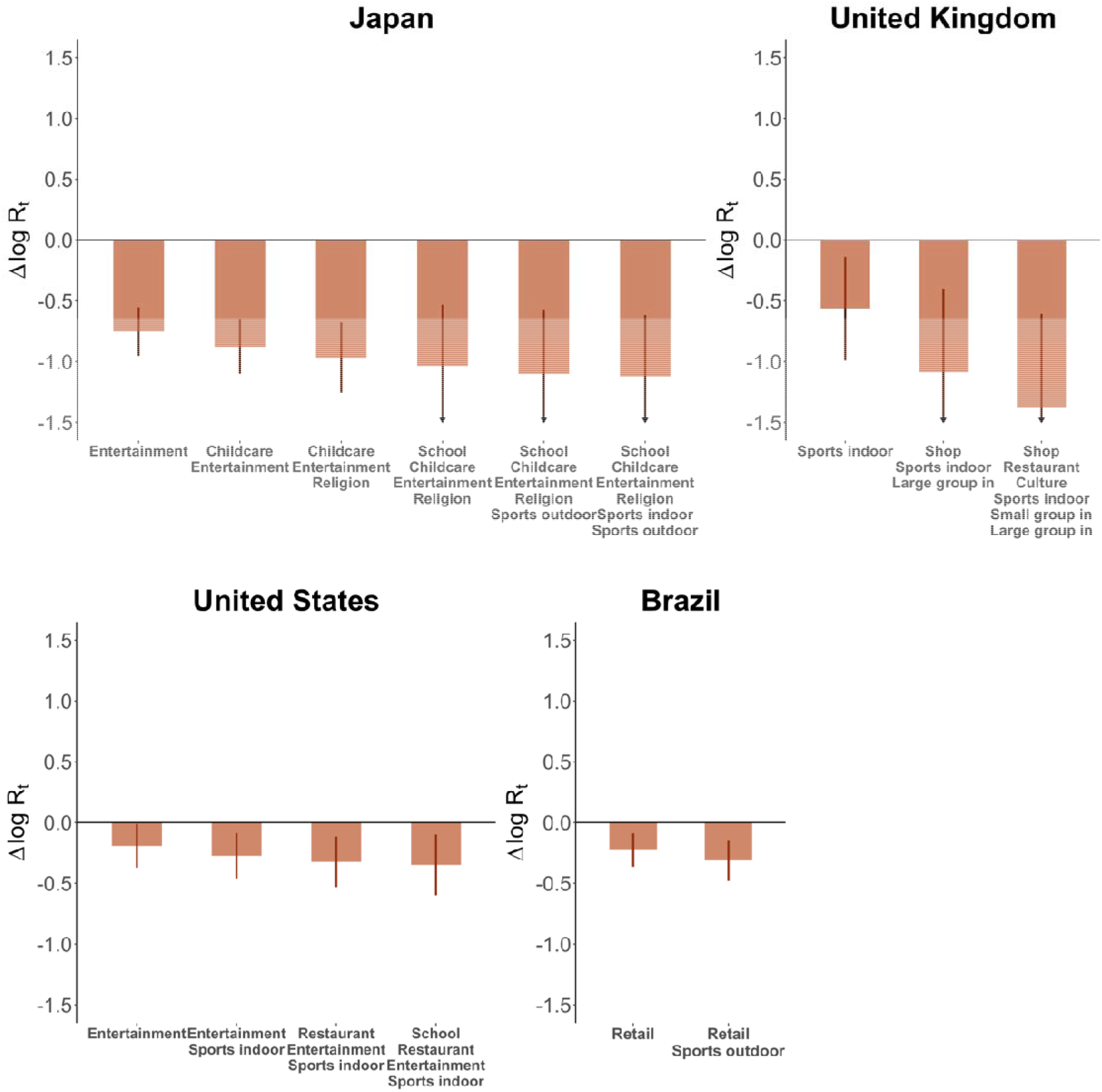
Maximum joint impacts of closing a number of activity spaces. We show the maximum impacts that could be achieved by closing a given number of types of activity spaces, till the maximum joint impacts are produced.

The combinations that generate the maximum effects are: closing schools, childcare centers, entertainment, religious, indoor sports venues and outdoor sports grounds in Japan (67%, 46 ∼ 80%); closing non-essential retails, restaurants, cultural venues and indoor sports venues in the United Kingdom (plus banning indoor gatherings whose effect is inseparable, 75%, 45 ∼ 88%); closing schools, restaurants, entertainment and indoor sports venues in the United States (29%, 9 ∼ 45%); and non-essential retails and outdoor sports grounds (27%, 13 ∼ 38%) in Brazil. Closing these sets of activity spaces could bring *R*_*t*_ below 1 for previous *R*_*t*_ up to 3.0 (1.9 ∼ 5.0) in Japan, 4.0 (1.8 ∼ 8.3) in the United Kingdom, 1.4 (1.1 ∼ 1.8) in the United States and 1.4 (1.1 ∼ 1.6) in Brazil. Note that the few activity space closures not satisfying the parallel trend assumption are excluded from this joint effect analysis since their impact estimates are not reliable (Table S3), but they may actually be able to further contribute to the reduction of *R*_*t*_.

### Impact of macro-level settlement characteristics

The results on the relationship between settlements’ population size and density and the fixed effects of spatial units are fairly consistent across the four countries (Table 1). Population size is negatively correlated with spatial unit’s fixed effect on *R*_*t*_ and this is statistically significant in three of the four countries, where the effect size ranges between 2.0% (1.1 ∼ 3.2%) to 4.9% (2.3 ∼ 7.5%) reduction of *R*_*t*_ per one million increase of population. The impact of density is less clear, yet none of the estimates is positively significant as suggested by the common beliefs mentioned in the Introduction. These results contradict the impression that large and densely populated cities tend to be epicenters and suggest that in terms of the reproduction number, large and dense cities are not riskier, but even less. Explanations for the negative relationship between settlement size and *R*_*t*_ might include better health infrastructures in large cities and people’s stronger awareness of the risk thus more cautious behavior (*24, 25*).

**Table 1.** Settlement characteristics on the fixed effects of spatial units.

| Characteristics | Japan | United Kingdom | United States | Brazil |
| --- | --- | --- | --- | --- |
| Size | -0.118<br>(0.113) | -0.029**<br>(0.0111) | -0.0219***<br>(0.00531) | -<br>0.0506***<br>(0.0139) |
| Density | -0.203<br>(0.392) | -0.00108<br>(0.00539) | 0.00453<br>(0.0201) | -0.0184**<br>(0.00648) |
| Proportion of elderly population | -0.102<br>(2.29) | -0.0325<br>(0.2) | 0.535***<br>(0.155) | -0.228<br>(0.387) |
| Proportion of Black | - | -0.105<br>(0.425) | -0.000726<br>(0.000542) | - |
| Proportion of Asian | - | 0.162<br>(0.124) | 0.00304*<br>(0.00148) | - |
| Personal income | -0.125<br>(0.105) | - | -<br>0.00686***<br>(0.000926) | 0.105*<br>(0.0435) |
| GDP per capita | 0.00494<br>(0.00855) | -0.00123*<br>(0.000592) | 0.002**<br>(0.000635) | -0.000228<br>(0.000374) |
| (Intercept) | 0.781<br>(1.68) | -0.158**<br>(0.0514) | 0.101**<br>(0.0386) | 0.217***<br>(0.0463) |
| Observations | 44 | 234 | 307 | 319 |
| R-squared | 0.294 | 0.0622 | 0.345 | 0.125 |
| Adjusted R-squared | 0.198 | 0.0373 | 0.329 | 0.111 |

\*p<0.05, \*\*p<0.01, \*\*\*p<0.001
Note: Results in this table are based on samples excluding outliers (Shimane in Japan, Mendip in the United Kingdom, and Indianapolis-Carmel-Anderson, Pittsfield and San Angelo in the United States), so that the residuals are normally distributed (Shapiro-Wilk test $p>0.05$ ). Results on full samples are very similar (shown in Table S4). The variance inflation factors are all below 7.

### Interaction between settlement characteristics and activity space closures

We also examine the interaction between macro-level settlement characteristics and activity space closures, to investigate whether the impacts of closing activity spaces would differ across different types of settlements. To do this, we re-estimate the maximum joint effects of activity space closures on separate samples of relatively large and small population, and separate samples of relatively high and low density. The high/low samples are split by the median population size (174,980 people) and density (681 people per squared kilometer) of all sample spatial units, except for Japan where the population size and density are generally much higher so that we use the median of its own (314,082 people and 5,671 people per squared kilometer respectively).

The comparisons are remarkably consistent across the four countries in terms of the interaction with settlement size, that the impacts of activity space closures are larger in relatively small settlements (Fig. 4A). The differences in the reductions of *R*_*t*_ by closing activity spaces range between 3% and 18%. The impacts are also larger in relatively low-density settlements except for in the United Kingdom and United States where the impacts are close to each other. The disparity in effect size reflects different share of infections accounted for by the specific activity spaces in different types of settlements, suggesting that a higher proportion of virus transmission in large settlements is related to places other than the specific activity spaces, which might include public transit, streets and other public spaces (*26*).

**Fig. 4.**
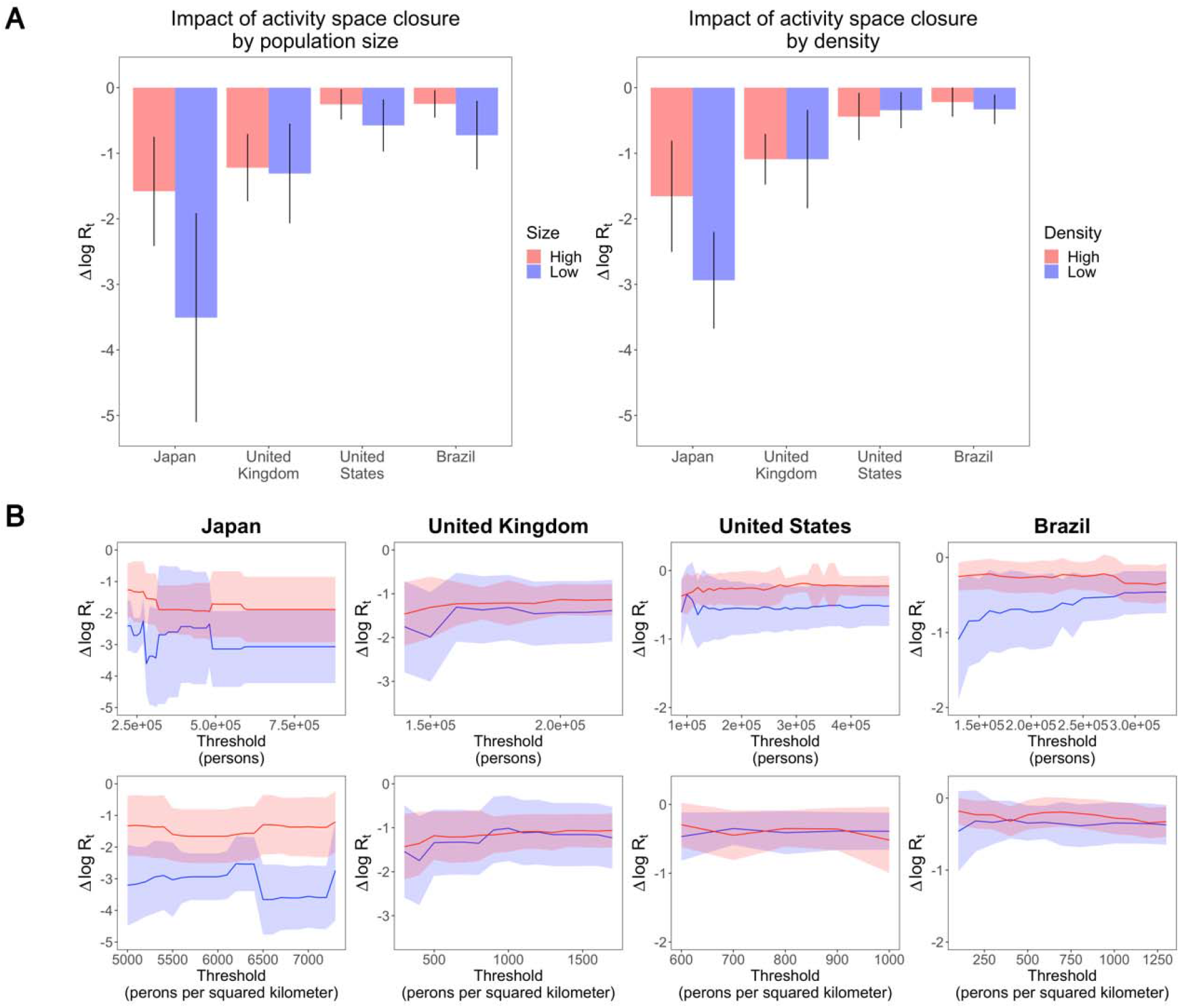
Impact of activity space closures in settlements with different population size and density. The error bars and ribbons indicate 95% confidence intervals.

To test whether the results hold when the population and density thresholds change, we repeat the analysis with a series of cut-off values between the first and third quantiles of population size and density in each country. The results are generally stable regardless of the threshold used to split the samples, and are particularly consistent in terms of settlement size: the effect sizes in relatively small settlements are always bigger than in large settlements in all sample countries (Fig. 4B). Similar pattern also exists with regard to density in Japan and less prominently in Brazil.

## Discussion

Our analysis quantifies the heterogenous risks of virus transmission associated with different activity spaces and settlements using evidences from the COVID-19 pandemic, and takes the first steps towards building systematic knowledge on the epistemological risks of places at multiple scales. The results demonstrate that governments could more cost-effectively contain the pandemic by closing a relatively small set of non-essential activity spaces, and such interventions would be more effective in relatively small settlements. Besides, contrary to common beliefs, settlements with relatively large population and density are not associated with higher risks of virus transmission. To our knowledge, this study is the first to attempt to estimate the impacts of closing individual types of activity spaces and to identify the varying effects of activity space closures in different types of settlements.

Though the risk of virus transmission at different types of activity spaces can also be evaluated with mechanistic modelling (*27, 28*), actual human behavior could be more complicated than experimental settings and our work provides data-driven evaluations. Our results demonstrate that closing a selected set of activity spaces could reduce *R*_*t*_ by 27-75% in our sample countries, without imposing a full lock down. Actually, the stay-at-home order does not demonstrate a statistically significant impact in reducing *R*_*t*_ in three of the four countries after controlling for other interventions and day and unit fixed effects (Table S2). This contradicts some previous findings, which however either do not rule out simultaneous voluntary behavioral changes or omit certain confounders (*14, 15, 19*). The magnitudes of the impacts are heterogeneous across countries, which could be affected by how people behave and interact at relevant places, the socioeconomic profile of the visitors, the physical conditions of relevant facilities, the intensity of intervention enforcement and so on. For example, the small effect size in the United States and Brazil might be explained by the relatively loose enforcement and non-compliance (*29, 30*). The heterogeneous results suggest that while some previous works seek to derive general conclusions on the effectiveness of interventions across countries (*8, 10*), such conclusions could run the risk of over-simplification and be misleading for policy making in individual countries given the factors mentioned above. Despite of the heterogeneity, it is common in the four sample countries that the closures of essential places including schools, childcare centers and offices do not demonstrate statistically significant effects in reducing *R*_*t*_ and non-essential activity spaces tend to be more effective, suggesting that governments could consider closing non-essential activity spaces as cost-effective interventions.

At the macro-level, our findings on the impacts of settlement size and density on *R*_*t*_ contradict the common belief that large and densely populated cities are more vulnerable to infectious disease (*31*). This could either because the seemingly increased connectivity and proximity among people do not actually enhance the chance for one infected person to transmit the virus to others, or such effect does exist but is offset by other positive factors of large and dense cities such as more healthcare resources driven by economy of scale and more cautious behavior of people. The exact causal chain could also involve the demography, economy even partisanship in different types of settlements (*25, 32*), which is subject to further study. Either way, these results lend more confidence to encouraging agglomeration of people and high-density development, as in the end, they are not associated with higher epistemological risks.

The finding that specific activity spaces account for a smaller proportion of transmission in relatively large settlements suggests that the pattern of virus transmission in these settlements is more complicated, which might be related to longer travel, more contacts on public transit and streets, home transmission in crowded residences and so on. It indicates that governments might need to take extra measures other than locking down to contain the pandemic in large cities, such as contact tracing or providing assistance to those living in poor conditions.

There are a number of limitations associated with our methodology. In terms of the causal identification strategy, the DiD method requires both parallel trend and exogeneity of the treatment. While the parallel trend assumption is examined with an event-study design, the exogeneity assumption could be challenged by unobserved confounders that affect both *R*_*t*_ and activity space closures. Though we are able to rule out a number of confounders by including a large set of interventions as well as unit and day fixed effects, there could still be endogeneity arising from omitted unit-specific time-varying factors. For instance, a sudden outburst of cases in a hotspot may affect both governments’ interventions and local residents’ cautionary behavior which then affects *R*_*t*_.

Second, since the impacts of closing different types of activity spaces are estimated in one model, the results could be subject to the so-called “table 2 fallacy” which refers to that the coefficients of confounders in a model are wrongly interpreted as full causal effects while they are actually only the direct effects (*33*). This problem applies if decisions to close activity spaces affect each other so that they become confounders. While this is possible, we suppose such relationship should be weak since the decision to close or reopen activity spaces tend to be more directly affected by the trends of infections, instead of the status of each other.

Third, we assume linear relationship between *R*_*t*_ and the independent variables in the entire analysis, which is a convenient assumption made by many studies on intervention effects in COVID-19 (*8, 10, 12, 16, 18*). However, the impact of closing one type of activity space may rely on the status of other activity spaces, since the corresponding activities could be complementary or substitutive to each other, leading to interacting effects. It is encouraging that studies which examine nonlinear relationships and sequence of interventions do not find significant patterns (*8, 19, 34*), but the issue cannot be fully closed.

By evaluating the infection risks of places, this work contributes to an emerging literature on the resilience and health of cities (*35-37*), as cities have become the dominant form of human settlement. Actually, health concerns have been a key factor in shaping the planning and policy in cities as early as the time of John Snow and Ebenezer Howard at the advent of modern cities. Our findings suggest that with increased human agglomeration and interaction, controlling epidemics is no longer only about confined areas such as hospitals, residences or the water supply system, but also the entire urban space. Understanding the linkage between places, human activities and diseases would be important for long- and short-term policy making in public health, urban planning, urban economy and other relevant fields.

## Materials and Methods

### Data

We curate a data set combining daily infection cases, government interventions (including activity space closures, stay-at-home orders and gathering bans) and the spatial, demographic and economic characteristics of the spatial units in our study, from the onset of the pandemic till August 15 2020. The infection case data are sourced from Japan Broadcasting Corporation’s case reports, the UK government, Johns Hopkins University and the Brazilian Ministry of Health. The timetable of government interventions is manually collected from the websites of national and state-level governments, which are the main levels of government making decisions on interventions. The settlement-related information is gathered from a number of official websites. More details on data sources are provided in Section S1.

### Estimating impacts of closing individual types of activity spaces

The causal impacts of closing individual types of activity spaces across all spatial units and subgroups of spatial units in a country are estimated with a two-way fixed effect model specified as follows

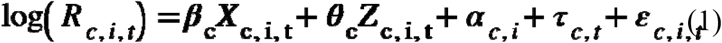

where *log(R*_*c,i,t*_) is the log-transformed instantaneous reproduction number in unit *i* of country *c* on day *t*; ***X***_***c***,***i***,***t***_ is a vector denoting the status of the eleven types of activity spaces and *β*_c_ denotes the corresponding coefficients to estimate. We log-transform *R*_*c,i,t*_ following the practice of relevant works (*10, 14*), based on the plausible assumption that the reduction of *R*_*c,i,t*_ by the closure of activity spaces should be proportional to the proportion of contacts avoided instead of an absolute value, and the impacts should be smaller when *R*_*c,i,t*_ is already low. *Z*_*c,i,t*_ and *θ*_*c*_ denote the status of five other government interventions and their coefficients (detailed description of these interventions in Table S1); *α*_*c,i*_ and *τ*_*c,t*_ denote the unit and time fixed effects, respectively; and *ε*_*c,i,t*_ denotes the error term. For the uncertainty over the parameters, we estimate robust standard errors allowing for *ε*_*c,i,t*_ to cluster at the unit level, to account for heterogeneity in the treatment effects (*38*).

### Estimating joint impacts of closing multiple types of activity spaces

The point estimates of the joint impacts are computed by summing the corresponding coefficients estimated by Eq. 1: 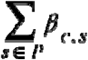, where *β*_*c,s*_ denotes the coefficient of closing activity space *s* in country *c* and *P* denotes a set of activity spaces. The standard errors are computed from the robust standard errors and covariances as follows

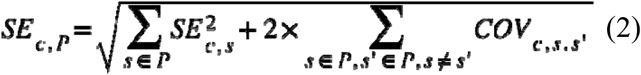

where *SE*_*c,P*_ denotes the standard error of the joint impacts of set *P* in country *c*; *SE*_*c,s*_ denotes the robust standard error of closing activity space *s* estimated by Eq. 1; and *COV*_*c,s,s’*_ is the covariance between the impacts of activity space *s* and *s’*.

### Estimating impacts of macro-level settlement characteristics

We take the unit fixed effects estimated by Eq. 1, which can be interpreted as the intrinsic reproduction number in each spatial unit, and model their relationship with the size and density of settlements while controlling for the proportion of elder population (over 65 or 60 years old depending on data availability), proportion of Black and Asian (in the United Kingdom and the United States only), the average income of residents and the per capita gross domestic product, using simple linear regression.

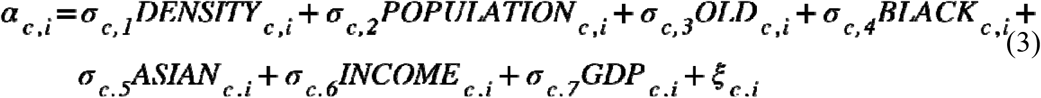

where *DENSITY*_*c,i*_, *POPULATION*_*c,i*_, *OLD*_*c,i*_, *BLACK*_*c,i*_, *ASIAN*_*c,i*_, *INCOME*_*c,i*_, *GDP*_*c,i*_ denote the density, population size, proportion of elder population, proportion of Black, proportion of Asian, residents’ income and per capita gross domestic product in unit *i*; σ_c,1_ to σ_c,7_ are their coefficients; *ξ*_*c,i*_ is the error term.

## Data Availability

Code and data are available on the following Github repository: https://github.com/lunliu454/infect_place. This work is licensed under a Creative Commons Attribution 4.0 International (CC BY 4.0) license, which permits unrestricted use, distribution, and reproduction in any medium, provided the original work is properly cited. To view a copy of this license, visit https://creativecommons.org/licenses/by/4.0/.

## Acknowledgments

We thank the High-performance Computing Platform of Peking University for providing the computation resource.

## Funding

Beijing Social Science Foundation 20GLA003 (LL)

National Natural Science Foundation of China 52008005 (LL)

Institute of Public Governance, Peking University YQZX202005 (LL)

## Author contributions

Conceptualization: LL, HW

Methodology: LL, HW, ZZ

Investigation: ZZ, WZ, TL, CC, YW, DN, CT, OA

Visualization: LL, ZZ

Writing—original draft: LL

Writing—review & editing: LL, HW, TL

