## Supplementary texts, figures and tables for "Infectiousness of places: The impact of human settlement and activity space in the transmission of COVID-19"

**This PDF file includes:**

Supplementary Text

Figs. S1 to S7

Tables S1 to S5

References (39 to 63)

Supplementary Text

S1 Data

We gathered data on demographic and economic characteristics of major large settlements, COVID-19 infections, and dates of place closures and other interventions in the selected settlements from a number of sources in the four sample countries. Detailed descriptions are provided below.

S1.1 Japan

**Spatial units and characteristic data.** The COVID-19 infection data can only be consistently acquired at the prefecture level in Japan, which is the first-level administrative division, therefore we can only use the prefectures as the spatial units of analysis (*21*). Prefectures can be much larger than individual continuous built-up areas, which is considered as a human settlement in this study, and usually contain more than one such area. In the main analysis, we use the population and density in the densely inhabited area of the largest city in a prefecture to represent the settlement conditions, since they reflect the conditions of the most populated area of the prefecture. However, our analysis shows that the conclusions are consistent when we use other indicators to represent the human settlements characteristics of the prefectures, including the total population of the largest city in a prefecture, as well as the total population and average density of all densely inhabited areas across a prefecture (Section S3.3). The population and land area of prefectures and cities as well as those of the densely inhabited areas are acquired from the portal site of official statistics of Japan, from where we also obtain the population age profile and household income in the prefectures (all in 2019, which is the latest data) (*39-41*). In addition, we obtain the gross domestic product in the prefectures in 2017 from the data portal of the Organization for Economic Co-operation and Development (*42*).

**COVID-19 infection data.** The infection case data in Japan is downloaded from the Github repository ‘covid19japan-data’, which is sourced from the Ministry of Health Labor and Welfare, the Japan Broadcasting Cooperation and prefectural governments (*21*). City-level data is only provided for a few major ones, thus invalid for our analysis.

**Place closure and other interventions.** The dates for closing and reopening various types of places and implementing other interventions are manually collected from the official websites of prefectural governments. To ensure accuracy of information, the data is collected by one person and double-checked by another. This work flow also applies to the data collection in other countries. Two pairs of interventions switched on and off simultaneously in our study period in Japan (Kendall’s *τ*=1) and thus are combined, which are banning small-size indoor gatherings and outdoor gatherings, and banning large-size indoor gatherings and outdoor gatherings. Information on the closure of outdoor sports grounds is incomplete thus excluded from further analysis.

S1.2 United Kingdom

**Spatial units and characteristic data.** Although the Office for National Statistics of the United Kingdom delineates ‘built-up areas’, which is consistent with our concept of settlements, they cannot be linked with the infection case data. Because to compute the daily infection cases in ‘built-up areas’, infection case data at the Middle layer Super Output Areas (MSOA) is needed, which can then be aggregated to the ‘built-up area’ level. Though the U.K. government does publish case data at the MSOA level, only latest data can be downloaded from the official website, so that we cannot compile the historical daily infection cases in the ‘built-up areas’. Instead, we use the Local Authority Districts (LAD) as the spatial unit of analysis. There are 290 LADs with population larger than 100,000 in 2020; however, these include 32 London boroughs which are combined, resulting in 259 spatial units (*43*). The sources for other demographic and economic characteristics of LADs are: population age profile from the mid-2020 edition of population estimates (*43*), ethnicity profile from Census 2011 (*44*), and gross domestic product per head from the 2019 edition of regional gross domestic product estimates (*45*) (we do not find personal income data at the LAD level).

**COVID-19 infection data.** The U.K. government publishes COVID-19 infection case data in fine-grained units (*46*). We request the data at the lower tier local authority level and aggregate the case numbers to LADs and London.

**Place closure and other intervention data.** The government interventions in COVID-19 are made by the four countries in UK, thus are collected from the official websites of the corresponding governments except for Northern Ireland, for which the information is not complete. Since the interventions are mostly uniform within the countries, four groups of interventions are highly simultaneous (Kendall’s *τ* >0.95) thus combined: closing non-essential retail and banning large-size indoor gatherings; closing restaurants, cultural venues and banning small-size indoor gatherings; closing entertainment venues and outdoor sports grounds; and banning small-size and large-size indoor gatherings. After cleaning missing information, we end up with 234 LAD spatial units.

S1.3 United States

**Spatial units and characteristic data.** We choose Metropolitan Statistical Areas (MSAs) as the spatial unit of analysis in the United States, which are regions with a relatively high population density at the core and socio-economically linked communities in the surroundings, delineated by the U.S. Office of Management and Budget. Since continuous built-up areas often spread beyond administrative borders, MSAs are more suitable for the analysis, although they are larger than a single continuous built-up area and contain rural areas in many cases. There are 363 MSAs with a population larger than 100,000 according to the 2019 estimates (*47*). To more accurately represent the population and density of major settlements in MSAs, we link MSAs with another statistical unit—urban areas, and compute the sum of urban population and urban land area in each MSA (*48*). The sources for other demographic and economic characteristics of MSAs are: population age profile from the 2010 Census (*49*), ethnicity profile from the American Community Survey (2019 data) (*50*), and personal income and gross domestic product per head from the Bureau of Economic Analysis (2019 and 2017 data, respectively) (*51, 52*).

**COVID-19 infection data.** The infection case data in the United States is downloaded from the COVID-19 Data Repository by the Center for Systems Science and Engineering (CSSE) at Johns Hopkins University, which publishes data at the county level (*53*). To acquire MSA level case data, we link MSAs with their containing counties using a look-up table provided by the Bureau of Economic Analysis and compute the total daily infections in the MSAs (*54*).

**Place closure and other intervention data.** We collect the dates of the interventions at the state level from the websites of state governments, which is the major level of government in charge of intervention policy making in COVID-19. By doing so, we do not account for the few cases in which county or local governments take alternative actions, which are relatively few (*34*). We do not find clear information on closing offices, outdoor sports grounds and banning indoor gatherings thus these interventions are not included in further analysis. We end up with 308 MSA spatial units after cleaning missing information.

S1.4 Brazil

**Spatial units and characteristic data.** We use the second-level administrative division—municipalities, as the units of study in Brazil. In 2020, there are 326 municipalities with more than 100,000 population in Brazil, which are taken as the subjects of analysis (*55*). According to Brazil’s last census, the proportion of urban population is larger than 80% in most of these municipalities (median = 96%), indicating that related demographic and economic statistics would mainly reflect the conditions of the dense urban settlements in the municipalities (*56*). The population age profile is also from the 2010 Census (*56*); the data for personal income (2019 data, available only at the state level) and gross domestic product per head is from the Brazilian Institute of Geography and Statistics (2018 data) (*57, 58*).

**COVID-19 infection data.** The infection case data in Brazil is downloaded from the Kaggle dataset ‘Coronavirus – Brazil’, which is sourced from the Coronavirus Brazil Dashboard published by the Brazilian Ministry of Health (*59, 60*). The data is available directly at the municipal level.

**Place closure and other intervention data.** The interventions are mainly decided by the state governments in Brazil, therefore collected from the state government websites. Three pairs of interventions are implemented and relaxed simultaneously during the study period (Kendall’s *τ* =1), which are the closures of schools and childcare centers, the bans on small-size indoor gatherings and small-size outdoor gatherings, and the bans on large-size indoor gatherings and large-size outdoor gatherings. We end up with 319 spatial units after cleaning missing information.

S1.5 Coding of place closure and other interventions

Table S1 describes the eleven types of place closures and five other interventions in this study. The interventions are coded 0, 0.5 or 1: 0 indicates that an intervention is not in place, 1 indicates a full restriction, and 0.5 indicates a partial restriction, including recommending instead of enforcing closures, or only closing establishments larger than a certain size, or shortening opening hours. Note that all interventions in Japan are non-compulsory, but since they are reported to have similar effects as full restrictions, we code them as 1 (*61*).

S2 Supplementary Method

S2.1 Estimating R_t_

The instantaneous reproduction number, *R_t_*, is the average number of secondary infected cases caused by a primary infected individual at time *t*, reflecting the speed of virus spread in a certain area, which is taken as the outcome of interest when analyzing the impacts of various types of places. *R_t_* is estimated using the method developed by Cori et al., which is widely used in epistemology (*62*). The method requires parameters of serial interval, which is set to be constant with mean=7.5 days and standard deviation=3.4 days following previous epidemiological investigation of COVID-19 (*63*). Further, to smooth out daily fluctuations, a 7-day sliding window is applied on the daily cases. Finally, *R_t_* estimates with a coefficient of variation larger than 0.3, indicating insufficient cases in the time window to generate reliable estimates, are excluded from further analysis.

S2.2 Testing for parallel pre-trend

To acquire reliable estimates from DiD analysis, the data need to meet the assumption of parallel trend, meaning that the outcome of interest should move in parallel trend in all units, absence of intervention. Since we cannot directly observe the counterfactual trend in units that have implemented a closure, we examine this assumption on the pre-intervention periods. This is implemented by an event study, adding terms indicating pre-intervention and post-intervention periods to the basic two-way fixed-effect model, specified as follows

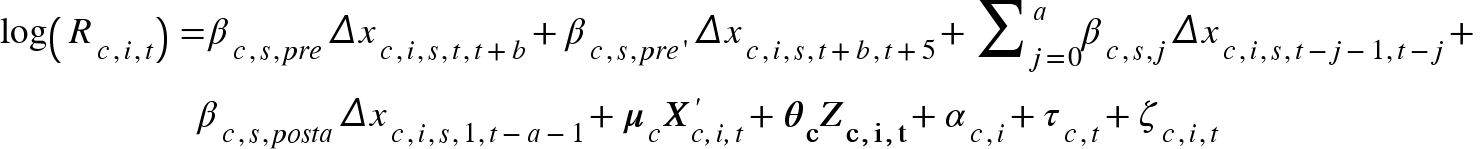
 (4)

where *Δx_c,i,s,t,t+b_* (*b* ≤ 5) denotes the change in the status of activity space *s* in unit *i* from day *t* to day *t+b* and the same applies to *Δx_c,s,i,t+b,t+5_*, *Δx_c,s,i,t-j-1,t-j_* and *Δx_c,s,i,1,t-a-1_*. Correspondingly, *β_c,s,pre_* denotes the estimate of pre-trend *b* days before a change, which is the coefficient of concern. *β_c,s,pre’_*, *β_c,s,j_* and *β_c,s,posta_* are to control for the effects in other periods: *β_c,s,pre’_* denotes the estimate of pre-trend *b* to 5 days before a change (if *b*=5, then the estimate is NA); *β_c,s,j_* is the effect *j* days (0 ≤ *j* ≤ a) after an intervention; *β_c,s,posta_* is the effect *a* days after a change, which should all be interpreted as compared to the period preceding to 5 days before a change. We test the pre-trend in 1 to 5 days before an intervention, since activity space closure and reopening tend to be quick decisions. The choice of *a* does not significantly affect the estimates, for which we choose 14, equaling to two weeks. ***X’_c,i,t_*** denotes the status of the other ten types of activity spaces in unit *i* on day *t* and ***μ_c_*** denotes the coefficients. The rest of the notations are the same as in Eq. 1. The inclusion of pre-intervention variable can be considered as a placebo test, which should be insignificant if the parallel trend assumption is satisfied.

S3 Robustness Check

To examine the robustness of the results, we conduct a series of experiments to test how alternative settings would affect our conclusion. First, we test whether the estimates are strongly affected by certain units in the data by re-running the models withholding part of the data. Second, we explore the confounding of possible missing variables. Last, we examine whether the interaction between activity space closures and settlement characteristics is sensitive to the indicators of settlement size and density.

S3.1 Sample selection

To test the sensitivity of our results to specific units, we re-estimate Eq. 1 by withholding one spatial unit at a time for *k* times, where *k* is the number of spatial units in each country. Figure S5 shows the distribution of point estimates in these alternative settings, which are mostly close to the estimates in the default setting. In the few cases where an alternative estimate departs from the default, the sign of the estimate does not change unless the default estimate is close to zero and not statistically significant.

S3.2 Missing variables

We control the status of five other interventions in estimating the impact of closing activity spaces. However, *R_t_* may also be influenced by other variables missed from our analysis. To explore whether our estimated impacts of closing activity spaces could be affected by missing variables, we assess (1) how the estimates change when we include additional confounding variables—in this case, requirements of wearing masks, and (2) how they change when we exclude existing variables. The results are shown in Figure S6, which demonstrates that the significance and magnitude of the estimates do not change much in these experimental conditions, lending more confidence to the robustness of our estimates to potentially missing variables.

S3.3 Alternative indicators of settlement characteristics

Since we use spatial units larger than continuous built-up settlements, there can be multiple indicators to represent the settlement conditions in a spatial unit. For example, the spatial unit in Japan—prefecture is the first-level administrative division and usually contains more than one large settlement. Statistics are available on the total population and land area of second-level divisions (cities, districts, villages), as well as those of densely inhabited areas in second-level divisions. In the main analysis, we use the population and density in the densely inhabited area of the largest city in a prefecture to represent the settlement conditions, since they reflect the conditions of the most populated area of the prefecture. However, there can be other plausible indicators, including the total population of the largest city in a prefecture and the total population of all densely inhabited areas across a prefecture as indicators of settlement size, and the average density of all densely inhabited areas across a prefecture as the indicator of settlement density. The spatial units analyzed in other countries are more similar to the extents of continuous built-up areas, but may also contain separate small settlements or non-built-up areas. We conduct similar robustness check on the United States, where we acquire data on both the population and density in the entire areas of MSAs and those in the urban areas. The latter are used in the main analysis and we test the sensitivity with the other indicators. Fig. S7 shows that the results are generally robust to the choice of indicators.

S4 Overview of the pandemic and interventions in the sample countries

S4.1 Japan

The first reported case of COVID-19 in Japan was confirmed on January 16 2020. The daily new cases were at two digits in February and March and rose to three digits in April. The central government started to declare state of emergency in April, which conferred relevant regional governments the power to implement partial or full lockdown. However, lockdown in Japan was distinct from many other countries in that government could only request residents to stay at home or businesses to close but could not force it, which relied on peer pressure and people’s deference to authority. On April 7 2020, the central government declared a state of emergency for seven prefectures and extended the emergency state to nationwide nine days later. From late March to mid-April, the 47 prefectures gradually requested stay-at-home and closure of non-essential facilities. Certain prefectures, such as Tokyo and Hokkaido, issued the requests even before the central government granted the power through the emergency declaration. Despite of the lenient approach, it was reported that there was fairly extensive voluntary compliance. The daily new cases fell back to two digits by mid-May, and remained at that low level till July. Correspondingly, prefectures started to release the interventions from late May, which however was followed by a stronger resurgence of infections in July and August (1,000+ new cases a day at the peak).

S4.2 United Kingdom

The first reported case of COVID-19 in the United Kingdom. was confirmed on January 31 2020. Cases grew mildly in February and then began to soar from early March with hundreds of new cases reported daily. Place-related interventions were launched from mid-March by the governments of the four countries of the United Kingdom, i.e. England, Wales, Scotland and Northern Ireland. The four countries have the autonomy to decide their own strategies but acted in a synchronized manner at the stage of locking down. All four countries closed schools on March 20 and three enforced stay-at-home order on March 26, followed by Northern Ireland on March 28. After reporting three to five thousand cases a day throughout April, infections gradually declined in May and June, and continued at a level of hundreds of new cases daily in July and August. The place-related interventions started to be lifted from the beginning of June and continued to be eased till the end of our study period except for in Leicester and Aberdeen City. The four countries acted less uniformly in lifting. For example, Wales continued to require people to stay within five miles from home till July 6, while people in England were free to leave their home from June 1.

S4.3 United States

The first reported case of COVID-19 in the United States was confirmed on January 20. The situation aggravated to thousands of new cases a day in March and more than 100,000 cases in total by late March. From March to April, the federal, state and local governments launched a number of orders and guidelines on social distancing, including ordering stay-at-home. Similar to Brazil, the power of ordering mass quarantine and closure lies primarily with the states, while the federal government provides recommendations and guidelines. Though all states imposed place-related interventions, the timetable and stringency varied a lot. Eight states did not mandate state-wide stay-at-home in our study period, while the dates of imposing stay-at-home in the other states ranged from March 19 to Apr 7. There were also up to more than 20 days differences in the closure of various businesses and facilities, if ever closed. In May and June, many states began to loosen the restrictions though cases were still increasing at a rate of more than 20,000 a day, which was followed by a resurgence to more than 60,000 cases a day in July. By the end of our study period, the United States became the country with the highest number of cases in the world.

S4.4 Brazil

The first reported case of COVID-19 in Brazil was confirmed on February 25, 2020, which was also the first confirmed case in the South America. The number of infections rose up to more than 1000 by mid-to-late March, when state and municipal governments started to declare an emergency situation and take measures such as closing non-essential businesses and suspending public activities. However, the interventions launched by state and local governments were undermined by the federal government especially the president, who kept dismissing the danger of the pandemic even issuing orders to expand the classification of essential businesses. Due to the autonomy of the states and the lack of federal-level coordination, heterogeneous interventions were observed across the country, e.g. the size of gathering banned by state governments in March varied from five people to hundreds.

Despite of the measures, the number of infections did not turn to a downward trajectory for months. By May 2020, Brazil became a new epicenter with more than 10,000 new cases confirmed daily. At this stage, several state and local governments introduced a full lockdown, which however was soon lifted in June together with previous interventions while cases still surged. The number of reported cases exceeded one million by late June, making Brazil the second largest hot spot of the pandemic after the United States. By the end of our study period, there were finally signs of decline in the spread of the virus.

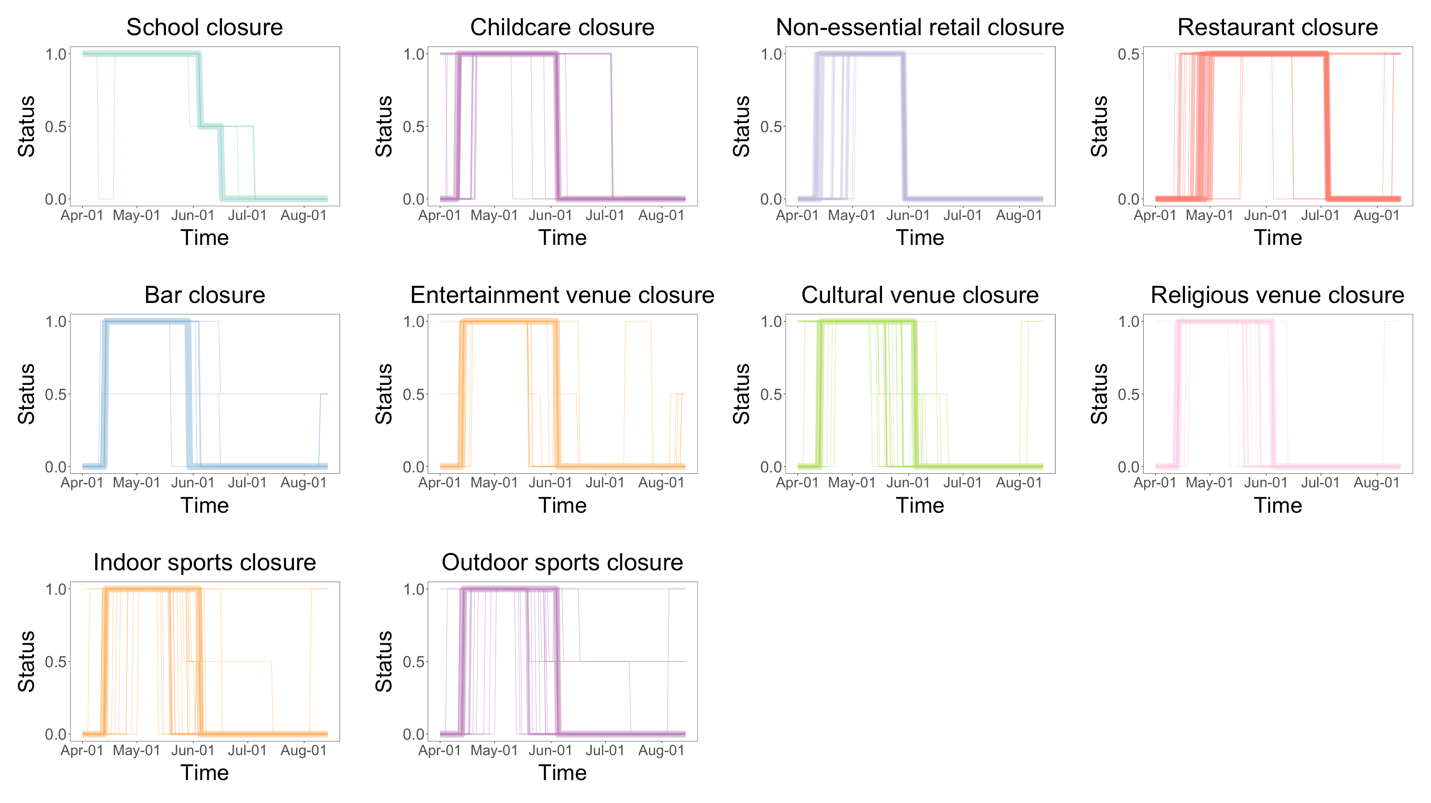

Fig. S1. Temporal dynamics of activity space closures and re-openings in Japan. The activity spaces switched between open (0), partly closed (0.5) and fully closed (1) during the study period (more details in Section S1.5). The widths of lines in the graphs are proportional to the number of spatial units in the corresponding status on the corresponding day.

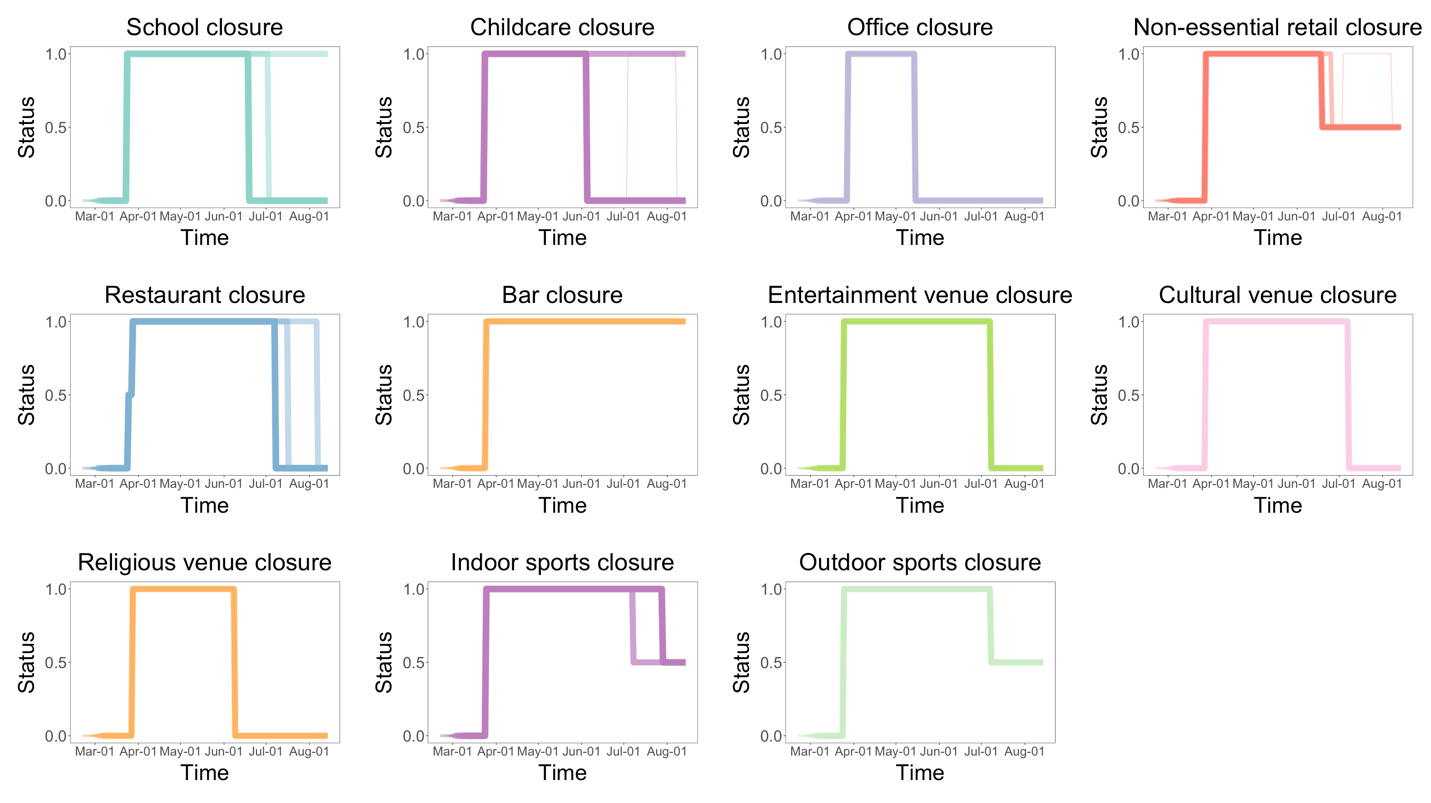

Fig. S2. Temporal dynamics of activity space closures and re-openings in the United Kingdom.

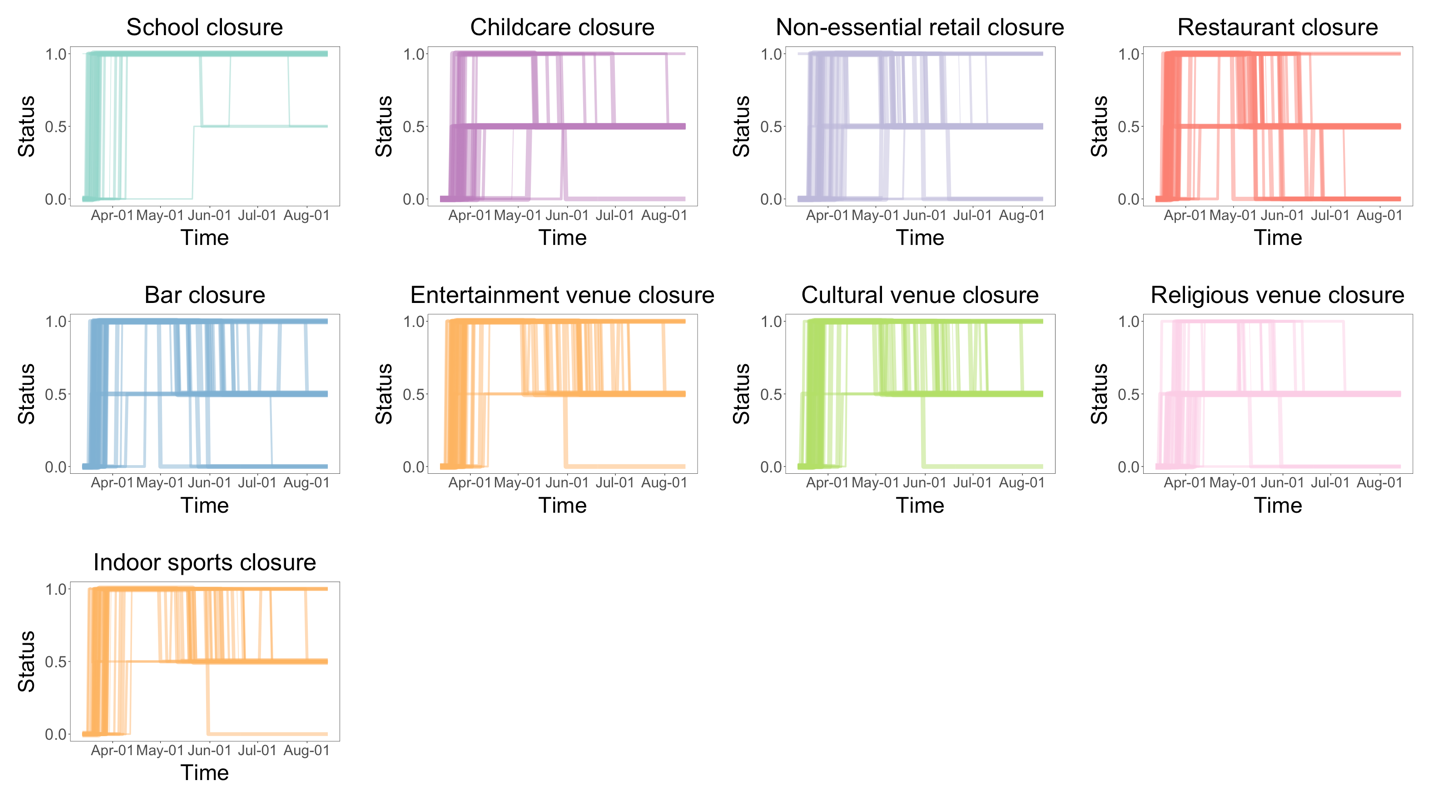

Fig. S3. Temporal dynamics of activity space closures and re-openings in the United States.

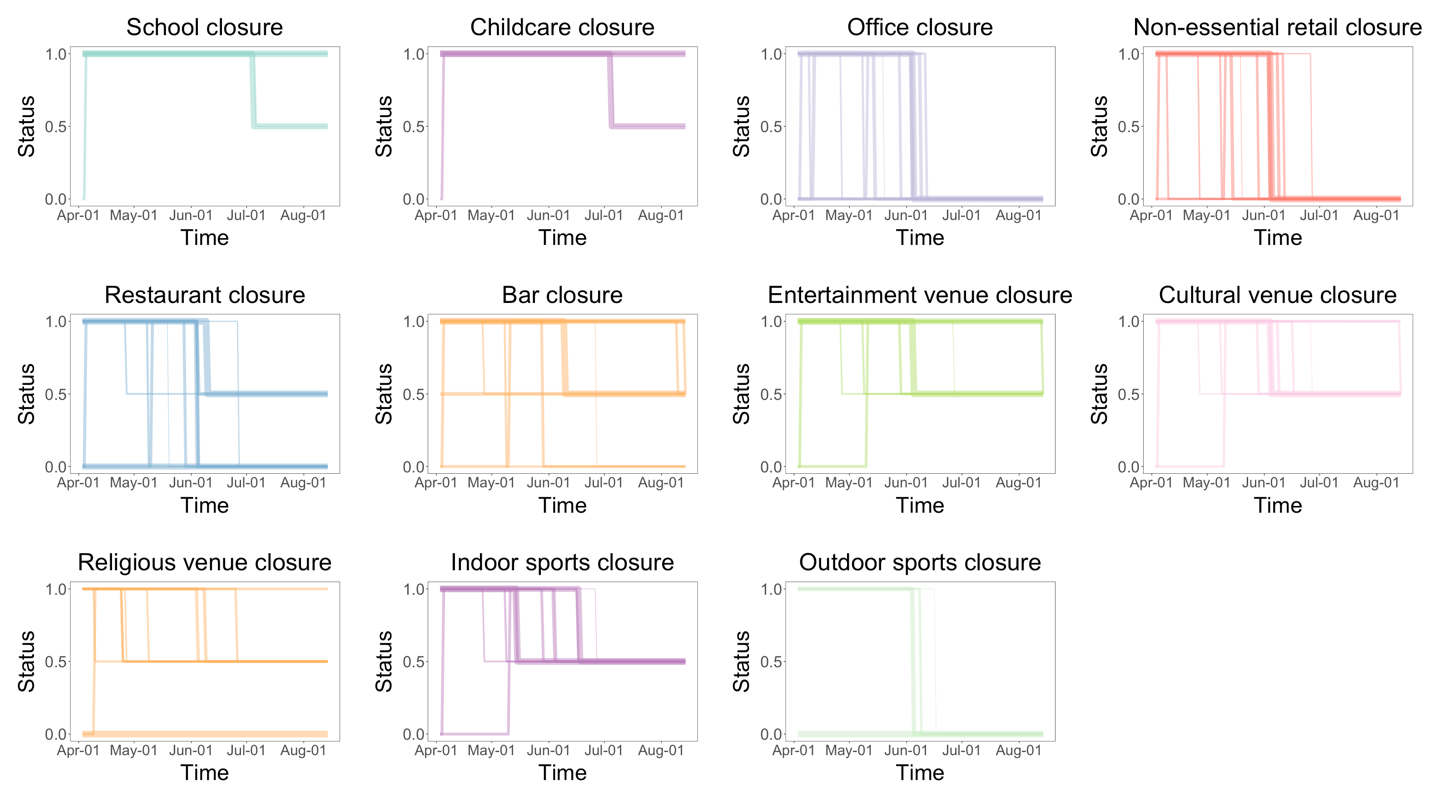

Fig. S4. Temporal dynamics of activity space closures and re-openings in Brazil.

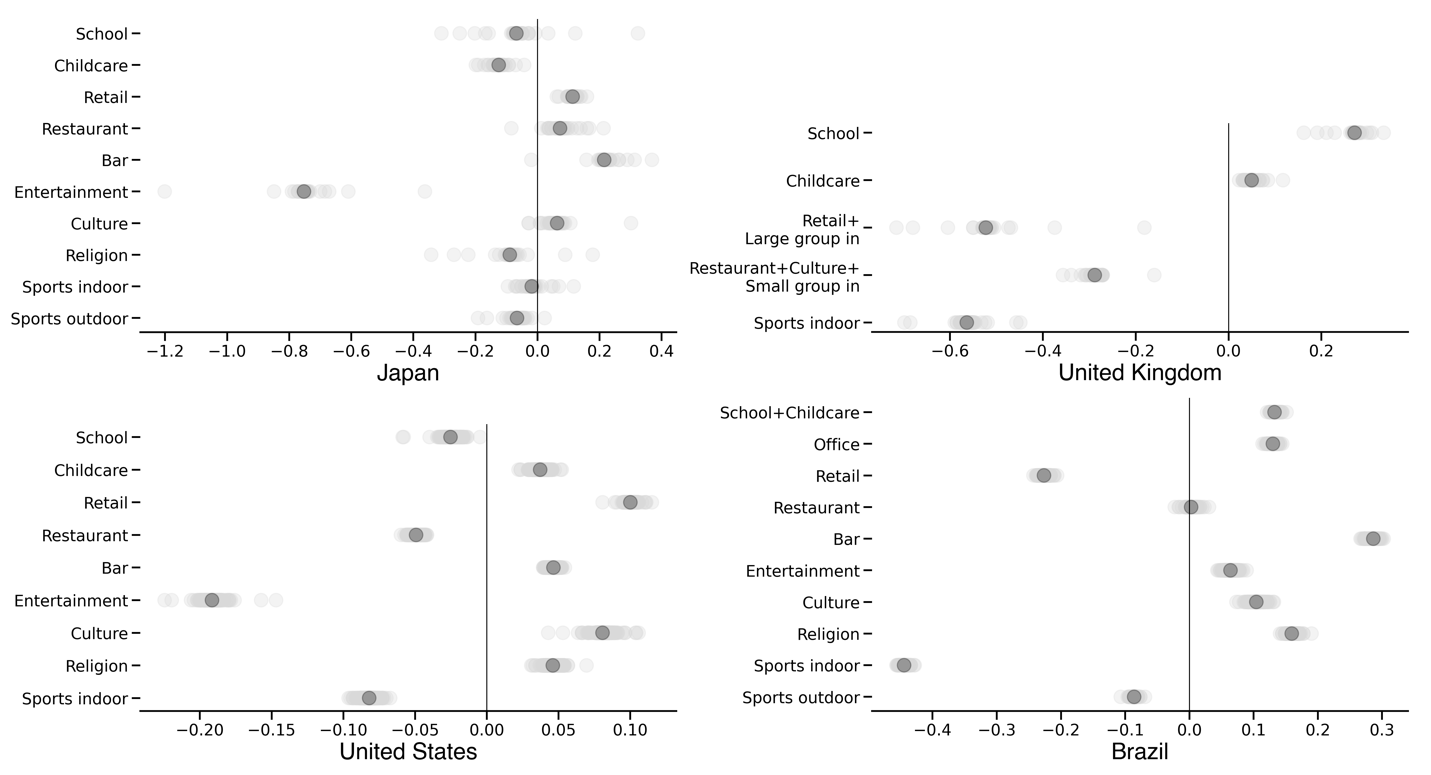

Fig. S5. Robustness of the estimates to withholding spatial units. Light grey dots are the alternative point estimates on the impacts of closing activity spaces when withholding one of the *k* spatial units in a country. Dark grey dots are the point estimates in the default setting, identical to the results in Fig. 2.

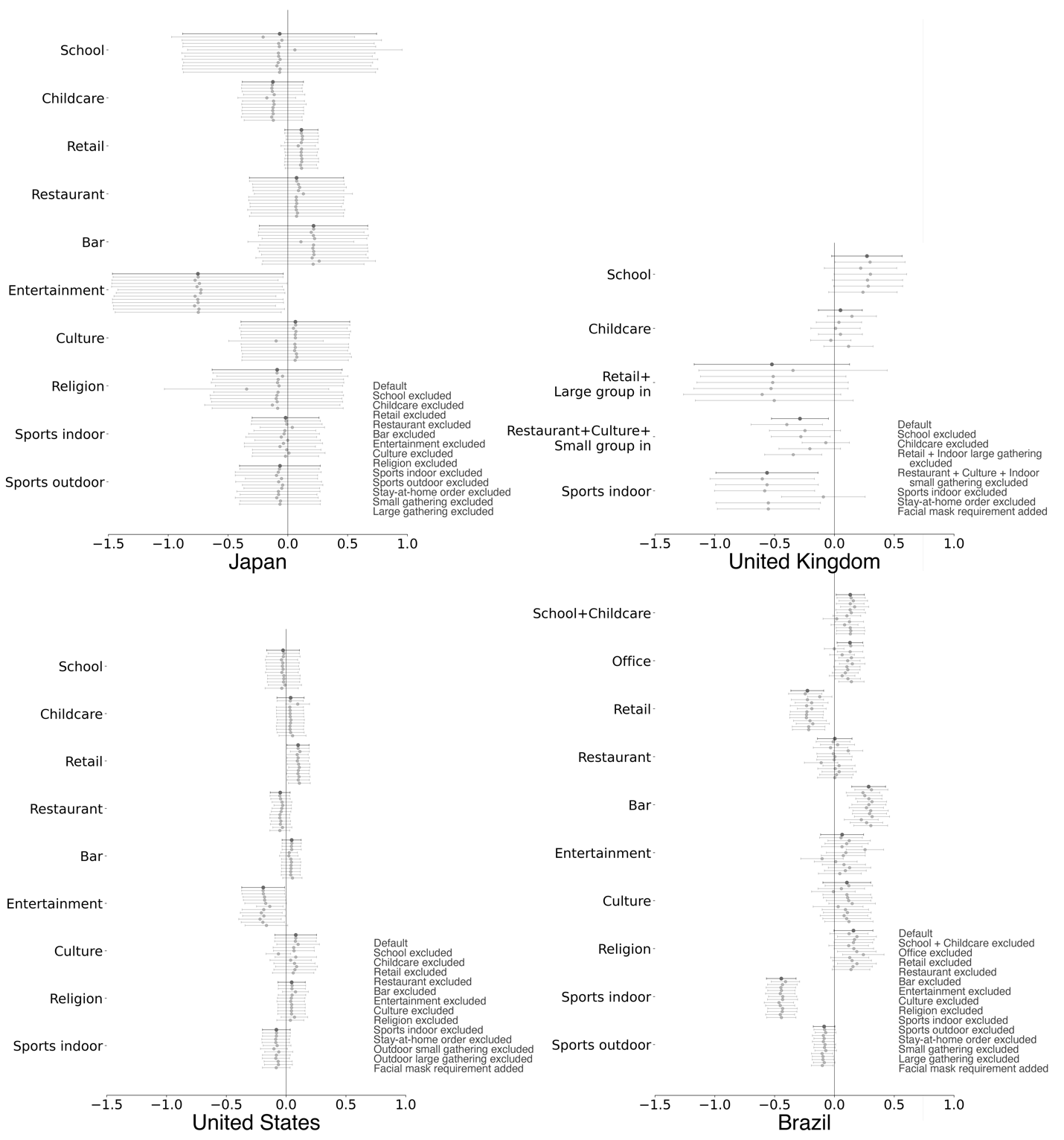

Fig. S6. Robustness of the estimates to omitting or including additional variables. The first bar in each group (dark grey) is the estimate in the default setting, followed by bars indicating estimates from alternative settings shown in the legend (except for the one that excludes the variable being examined).

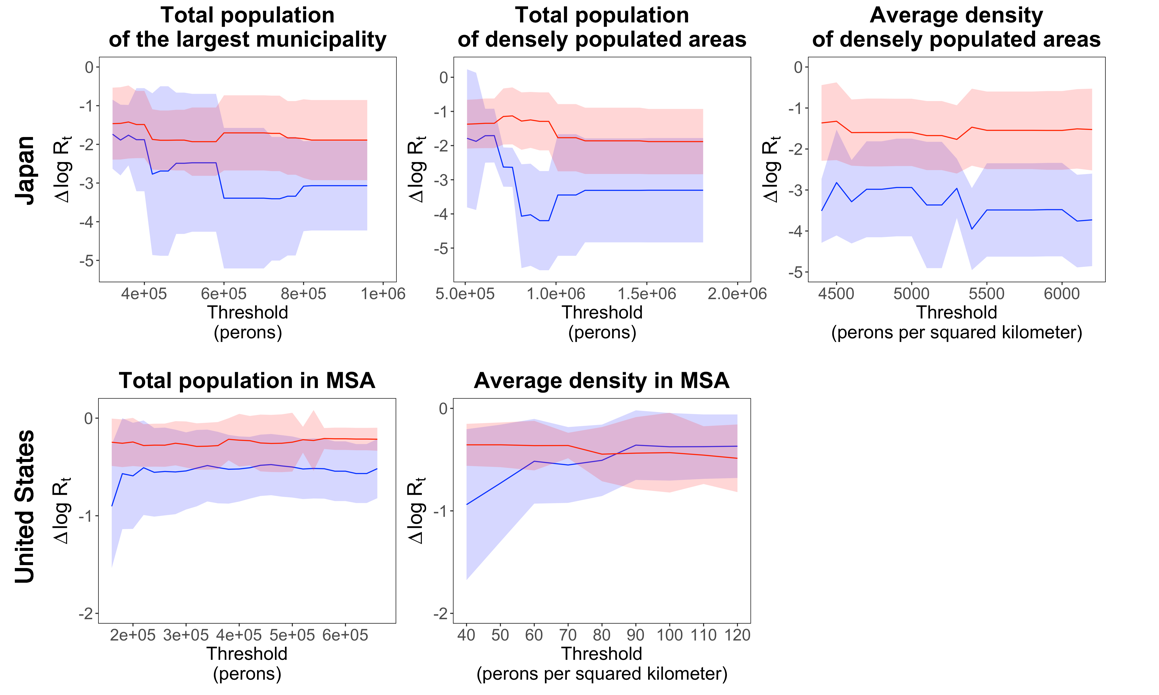

Fig. S7. Interaction between alternative indicators of settlement size and density and the impacts of activity space closures. The ribbons indicate 95% confidence intervals. The blue lines are estimates from low-value groups and red lines are from high-value groups.

Table S1. Place closures and other interventions.

| **Intervention** | **Notes** |
| --- | --- |
| ***Place closures*** |  |
| Closing schools | Including all types of schools. If face-to-face teaching is allowed only for certain types of schools or classes, then this is considered as a partial restriction. |
| Closing childcare centers | Note that the service for essential workers usually remain. |
| Closing offices |  |
| Closing non-essential retails | Including all retail establishments other than those considered essential, such as grocery stores, supermarkets and pharmacies. The definition of essential retails may differ across countries and regions. |
| Closing restaurants | Including restaurants, cafes, canteens, etc. Note that food delivery and take-away service usually remain. |
| Closing bars |  |
| Closing entertainment venues | Including clubs, discos, cinemas, karaoke, theaters, etc. |
| Closing cultural venues | Including museums, libraries, galleries, etc. |
| Closing religious venues | Including worship places for various religions. |
| Closing indoor sports venues | Including gyms, dance studios, indoor sports fields, etc. |
| Closing outdoor sports grounds | Including playgrounds, sports fields, etc. |
| ***Other interventions*** |  |
| Ordering stay-at-home | Requiring citizens to stay at home unless going out for essential purposes such as working in the essential sectors, shopping for necessities and seeing doctors. There can be small variations in full restriction in certain cases, for example people are allowed to travel within five miles from home in Wales in early June 2020, in which case the intervention is still coded as 1. |
| Banning small-size outdoor gatherings | The size limit is at most 10 people. |
| Banning large-size outdoor gatherings | The size limit is larger than 10. |
| Banning small-size indoor gatherings | The size limit is at most 10 people. |
| Banning large-size indoor gatherings | The size limit is larger than 10. |

Table S2. Full results of basic models.

| **Japan** | | **United Kingdom** | | **United States** | | **Brazil** | |
| --- | --- | --- | --- | --- | --- | --- | --- |
| **Intervention** | **Estimate** | **Intervention** | **Estimate** | **Intervention** | **Estimate** | **Intervention** | **Estimate** |
| ***Activity space closures*** | | | | | | | |
| School | -0.068  (0.414) | School | 0.271*  (0.150) | School | -0.0253  (0.070) | School+  Childcare | 0.133**  (0.060) |
| Childcare | -0.125  (0.131) | Childcare | 0.0498  (0.094) | Childcare | 0.0372  (0.058) | Office | 0.13**  (0.055) |
| Retail | 0.113  (0.071) | Retail+  Large group in | -0.523  (0.331) | Retail | 0.1**  (0.047) | Retail | -0.226***  (0.070) |
| Restaurant | 0.0727  (0.201) | Restaurant+  Culture+  Small group in | -0.288**  (0.122) | Restaurant | -0.0493  (0.042) | Restaurant | 0.00266  (0.074) |
| Bar | 0.215  (0.231) | Bar | NaN  (0.000) | Bar | 0.0465  (0.041) | Bar | 0.286***  (0.072) |
| Entertainment | -0.752**  (0.364) | Entertainment+  Sports outdoor | NaN  (0.000) | Entertainment | -0.192**  (0.092) | Entertainment | 0.064  (0.092) |
| Culture | 0.0634  (0.232) | Sports indoor | -0.564**  (0.218) | Culture | 0.0807  (0.089) | Culture | 0.104  (0.102) |
| Religion | -0.0892  (0.278) |  |  | Religion | 0.046  (0.058) | Religion | 0.159*  (0.083) |
| Sports indoor | -0.0184  (0.143) |  |  | Sports indoor | -0.082  (0.059) | Sports indoor | -0.444***  (0.063) |
| Sports outdoor | -0.0659  (0.172) |  |  |  |  | Sports outdoor | -0.086*  (0.046) |
| ***Other interventions*** | | | | | | | |
| Stay-at-home | -0.109  (0.178) | Stay-at-home | -0.236**  (0.116) | Stay-at-home | -0.0284  (0.028) | Stay-at-home | 0.118***  (0.041) |
| Small group in+out | 0.309  (0.263) |  |  | Small group out | -0.0458*  (0.025) | Small group in+  out | -0.347***  (0.098) |
| Large group in+out | 0.0566  (0.442) |  |  | Large group out | 0.0942**  (0.038) | Large group in+  out | 0.102  (0.100) |
| Observations | 2400 | 24285 |  | 41752 |  | 34812 |  |
| R-squared | 0.605 | 0.644 |  | 0.549 |  | 0.29 |  |
| Adjusted R-squared | 0.57 | 0.638 |  | 0.544 |  | 0.281 |  |

*p<0.1, **p<0.05, ***p<0.01

Table S3. Full results of pre-trend tests.

| **Intervention** | **1 day** | **2 days** | **3 days** | **4 days** | **5 days** | **Intervention** | **1 day** | **2 days** | **3 days** | **4 days** | **5 days** |
| --- | --- | --- | --- | --- | --- | --- | --- | --- | --- | --- | --- |
| **Japan** |  |  |  |  |  | **United Kingdom** | | | | | |
| School | 0.146  (0.599) | -0.0829  (0.568) | -0.239  (0.56) | -0.31  (0.557) | -0.375  (0.558) | School | **0.353****  **(0.131)** | **0.365****  **(0.131)** | **0.364****  **(0.126)** | **0.387****  **(0.12)** | **0.401*****  **(0.117)** |
| Childcare | -0.204  (0.165) | -0.197  (0.167) | -0.218  (0.159) | -0.196  (0.157) | -0.17  (0.151) | Childcare | **-0.33*****  **(0.0767)** | **-0.32*****  **(0.0722)** | **-0.32*****  **(0.07)** | **-0.27*****  **(0.0667)** | **-0.24*****  **(0.067)** |
| Retail | 0.125  (0.122) | 0.103  (0.117) | 0.107  (0.107) | 0.0881  (0.0954) | 0.0672  (0.0862) | Retail+  Large group in | 0.288  (0.257) | 0.279  (0.258) | 0.286  (0.243) | 0.333  (0.237) | 0.343  (0.233) |
| Restaurant | 0.215  (0.274) | 0.19  (0.246) | 0.21  (0.225) | 0.222  (0.213) | 0.211  (0.198) | Restaurant+  Culture+  Small group in | -0.0484  (0.132) | -0.0305  (0.118) | -0.0416  (0.108) | -0.0486  (0.0936) | -0.0596  (0.091) |
| Bar | -0.269  (0.333) | -0.257  (0.315) | -0.225  (0.286) | -0.207  (0.249) | -0.198  (0.225) | Sports indoor | 0.0645  (0.367) | 0.112  (0.351) | 0.187  (0.354) | 0.229  (0.338) | 0.251  (0.335) |
| Entertainment | -0.207  (0.156) | -0.162  (0.143) | -0.114  (0.132) | -0.042  (0.124) | 0.0116  (0.119) |  |  |  |  |  |  |
| Culture | -0.0457  (0.17) | -0.0521  (0.165) | -0.0405  (0.164) | -0.0030  (0.155) | 0.0224  (0.149) |  |  |  |  |  |  |
| Religion | 0.0125  (0.285) | -0.0014  (0.263) | 0.0272  (0.261) | 0.0734  (0.245) | 0.0913  (0.24) |  |  |  |  |  |  |
| Sports  indoor | -0.0099  (0.216) | -0.0441  (0.216) | -0.0402  (0.216) | -0.0164  (0.208) | -0.0016  (0.199) |  |  |  |  |  |  |
| Sports  outdoor | -0.0582  (0.227) | -0.082  (0.219) | -0.0784  (0.214) | -0.0556  (0.203) | -0.0452  (0.196) |  |  |  |  |  |  |
| **United States** | | | | | | **Brazil** |  |  |  |  |  |
| School | -0.084  (0.0746) | -0.0542  (0.0861) | -0.0313  (0.0929) | -0.0041  (0.102) | 0.0425  (0.112) | School+  Childcare | 0.132  (0.114) | 0.0363  (0.1) | -0.0202  (0.0971) | -0.0561  (0.0943) | -0.0755  (0.0901) |
| Childcare | -0.0708  (0.0519) | -0.0949  (0.0533) | **-0.112***  **(0.0551)** | -0.103  (0.0583) | -0.11  (0.0582) | Office | -0.0428  (0.0451) | -0.0399  (0.0425) | -0.0352  (0.0407) | -0.0299  (0.0387) | -0.0184  (0.0366) |
| Retail | 0.0139  (0.0323) | 0.0077  (0.0339) | 0.0026  (0.036) | 0.00991  (0.0383) | 0.0106  (0.0395) | Retail | -0.0691  (0.0479) | -0.0619  (0.0448) | -0.0527  (0.0424) | -0.0375  (0.0399) | -0.0181  (0.0372) |
| Restaurant | -0.04  (0.0327) | -0.0374  (0.0346) | -0.0388  (0.0361) | -0.037  (0.0379) | -0.0315  (0.0392) | Restaurant | -0.0829  (0.073) | -0.0774  (0.0696) | -0.0837  (0.0679) | -0.0817  (0.0653) | -0.0663  (0.0628) |
| Bar | -0.0322  (0.0367) | -0.0307  (0.0392) | -0.037  (0.041) | -0.0378  (0.0426) | -0.0324  (0.0436) | Bar | 0.0604  (0.0782) | 0.0539  (0.0764) | 0.0322  (0.0741) | 0.0201  (0.0693) | 0.0217  (0.0646) |
| Entertainment | **-0.081***  **(0.0369)** | **-0.080***  **(0.04)** | **-0.084***  **(0.0416)** | -0.084  (0.0433) | -0.0783  (0.045) | Entertainment | -0.117  (0.0943) | -0.135  (0.0914) | -0.161  (0.0873) | -0.155  (0.0825) | -0.149  (0.0766) |
| Culture | -0.021  (0.0351) | -0.0236  (0.0375) | -0.0241  (0.039) | -0.02  (0.0411) | -0.0109  (0.0428) | Culture | 0.00231  (0.0922) | 0.0242  (0.0858) | 0.0249  (0.0813) | 0.0463  (0.0766) | 0.0706  (0.0724) |
| Religion | -0.0554  (0.0464) | -0.0696  (0.0493) | -0.0918  (0.0506) | -0.102  (0.0521) | **-0.123***  **(0.053)** | Religion | 0.204  (0.11) | **0.239***  **(0.108)** | **0.238***  **(0.105)** | **0.217***  **(0.103)** | **0.204***  **(0.102)** |
| Sports  indoor | -0.0404  (0.0369) | -0.0299  (0.0397) | -0.0364  (0.041) | -0.0326  (0.043) | -0.028  (0.0447) | Sports  indoor | **-0.33*****  **(0.0749)** | **-0.36*****  **(0.0726)** | **-0.36*****  **(0.0695)** | **-0.34*****  **(0.0662)** | -**0.31*****  **(0.0619)** |
|  |  |  |  |  |  | Sports outdoor | -0.0212  (0.049) | -0.00321  (0.0466) | 0.00729  (0.0452) | 0.0237  (0.044) | 0.0353  (0.0429) |

*p<0.05, **p<0.01, ***p<0.001

Table S4. Settlement characteristics model results with full sample

| **Characteristics** | **Japan** | **United Kingdom** | **United States** | **Brazil** |
| --- | --- | --- | --- | --- |
| Size | -0.276  (0.141) | -0.0312**  (0.0118) | -0.021***  (0.00564) | -0.0506***  (0.0139) |
| Density | 0.326  (0.488) | -0.00251  (0.00574) | 0.00372  (0.0214) | -0.0184**  (0.00648) |
| Proportion of elderly population | 2.65  (2.88) | -0.0368  (0.213) | 0.448**  (0.165) | -0.228  (0.387) |
| Proportion of black | - | -0.0525  (0.453) | -0.000828  (0.000578) | - |
| Proportion of Asian | - | 0.157  (0.132) | 0.00298  (0.00157) | - |
| Personal income | -0.0174  (0.132) | - | -0.0068***  (0.000988) | 0.105*  (0.0435) |
| GDP per capita | 0.00841  (0.011) | -0.00133*  (0.000631) | 0.00179**  (0.000678) | -0.000228  (0.000374) |
| (Intercept) | -1.82  (2.07) | -0.158**  (0.0547) | 0.123**  (0.041) | 0.217***  (0.0463) |
| Observations | 44 | 234 | 307 | 319 |
| R-squared | 0.294 | 0.0637 | 0.315 | 0.125 |
| Adjusted R-squared | 0.201 | 0.039 | 0.299 | 0.111 |

*p<0.1, **p<0.05, ***p<0.01
